# CAN-NPI: A Curated Open Dataset of Canadian Non-Pharmaceutical Interventions in Response to the Global COVID-19 Pandemic

**DOI:** 10.1101/2020.04.17.20068460

**Authors:** Liam G. McCoy, Jonathan Smith, Kavya Anchuri, Isha Berry, Joanna Pineda, Vinyas Harish, Andrew T. Lam, Seung Eun Yi, Sophie Hu, COVID-19 Canada Open Data Working Group: Non-Pharmaceutical Interventions, Benjamin Fine

## Abstract

Non-pharmaceutical interventions (NPIs) have been the primary tool used by governments and organizations to mitigate the spread of the ongoing pandemic of COVID-19. Natural experiments are currently being conducted on the impact of these interventions, but most of these occur at the subnational level - data not available in early global datasets. We describe the rapid development of the first comprehensive, labelled dataset of 1640 NPIs implemented at federal, provincial/territorial and municipal levels in Canada to guide COVID-19 research. For each intervention, we provide: a) information on timing to aid in longitudinal evaluation, b) location to allow for robust spatial analyses, and c) classification based on intervention type and target population, including classification aligned with a previously developed measure of government response stringency. This initial dataset release (v1.0) spans January 1st, and March 31st, 2020; bi-weekly data updates to continue for the duration of the pandemic. This novel dataset enables robust, inter-jurisdictional comparisons of pandemic response, can serve as a model for other jurisdictions and can be linked with other information about case counts, transmission dynamics, health care utilization, mobility data and economic indicators to derive important insights regarding NPI impact.

## Background and Significance

Since the first case of COVID-19 was reported in Canada in January 2020, there have been a total of 30,968 reported cases and 1,271 reported deaths as of April 16, 2020^1^. In the absence of population immunity or an effective medical treatment, traditional public health interventions (e.g. physical distancing, testing, contact-tracing, and hand hygiene) are critical to protect population health^2,3^. These non-pharmaceutical interventions (NPIs) have been the primary tool employed by governments and organizations to reduce the spread of the virus, and avoid the possibility that peak case numbers overwhelm healthcare capacity^2,4,5^. In Canada, NPIs have included the closure of borders and bans on non-essential travel, as well as the imposition of voluntary or mandatory physical distancing measures.

While some NPI policies have been implemented at a national scale, much of the authority and responsibility to oversee rollout of these policies falls on provincial, territorial, and municipal governments^6^. As such, there has been substantial variability in what, when, and how NPIs have been implemented across Canada — highlighting the importance of a subnational lens of data gathering and analysis.

We present the first comprehensive, open dataset containing detailed information about all publicly available NPIs that have been implemented by governments and major private organizations in Canada in response to COVID-19. To ensure a comprehensive review that captures jurisdictional differences, we collect NPIs at the Canadian federal level, in all ten provinces and three territories, as well as in the twenty largest Census Metropolitan Areas (CMAs). Each intervention is labelled based on timing, location, intervention type, target population, and a previously developed measure of government response stringency.

The ultimate value of this dataset is to help researchers and policy-makers make sense of the first phase of NPIs, which are essentially a set of natural experiments occurring across the globe. The first of its kind in Canada, this dataset enables research characterizing the nature of Canada’s COVID-19 response at a regional level. Regional NPI data can now be linked with other information about case counts, transmission dynamics, health care utilization, mobility data and economic indicators to derive important insights regarding NPI impact. Moreover, each Canadian region can be compared to other Candian or international jurisdiction to facilitate comparative analyses. We hope this dataset can also serve as a model for subnational NPI data collection from other jurisdictions.

## Dataset Generation Methods

In this dataset, we define a non-pharmaceutical intervention (NPI) as any publicly-announced program, statement, enforceable order, initiative, or operational change originating from any public or private organization in response to COVID-19—whether to curtail its transmission or mitigate its social and economic ramifications. This includes distancing measures (including closures), infection control measures (excluding vaccination or medical treatment), testing strategies, public announcements, media campaigns, and social and fiscal measures, among others. Broadly, these are all parameters that influence the behaviour of individuals in an effort to limit the spread and economic impact of COVID-19^3^.

### Review Team Composition

Primary review was performed by the COVID-19 Canada Open Data Working Group: Non-Pharmaceutical Interventions, a 34-member team primarily composed of medical students and graduate students from the University of Toronto. This team was recruited one of the authors via a standardized onboarding process.

### Selection of sources

We identified NPIs implemented at all different levels of government, including the Canadian federal level, provincial and territorial level, and the municipal level for the 20 largest CMAs in Canada (see Supplementary Material, Table S1). We also included NPIs implemented by private organizations at these regional levels. For the first version of the dataset, we conducted a comprehensive scan of all COVID-19-related NPIs and changes to testing requirements or case definitions between January 1st and March 31st, 2020. Future versions will be updated every 2 weeks until the pandemic ends. Interventions were recorded only for the administrative level responsible for them, such that an intervention recorded at the provincial level was not repeated at the municipal level.

Articles were selected through a comprehensive online environmental scan, which is appropriate for the rapidly evolving nature of the ongoing pandemic and the variety of avenues through which information is announced^8^ (**Figure 1**). A hierarchy of preferred sources was used to identify and code interventions (Supplementary Material Table S4). First, official government sources were reviewed in full and any COVID-19-related announcements were identified as the gold standard for data inclusion. Government sources include press releases on the official websites of provincial and territorial governments, Ministries of Health, or Provincial Public Health Commissions. Second, additional information was identified using purposive search methods for COVID-19-related articles and online reports from accredited news agencies. Finally, we include updates provided by the official social media accounts of governmental or public health institutions — such as provincial Chief Medical Officers of Health.

**Figure 1.**
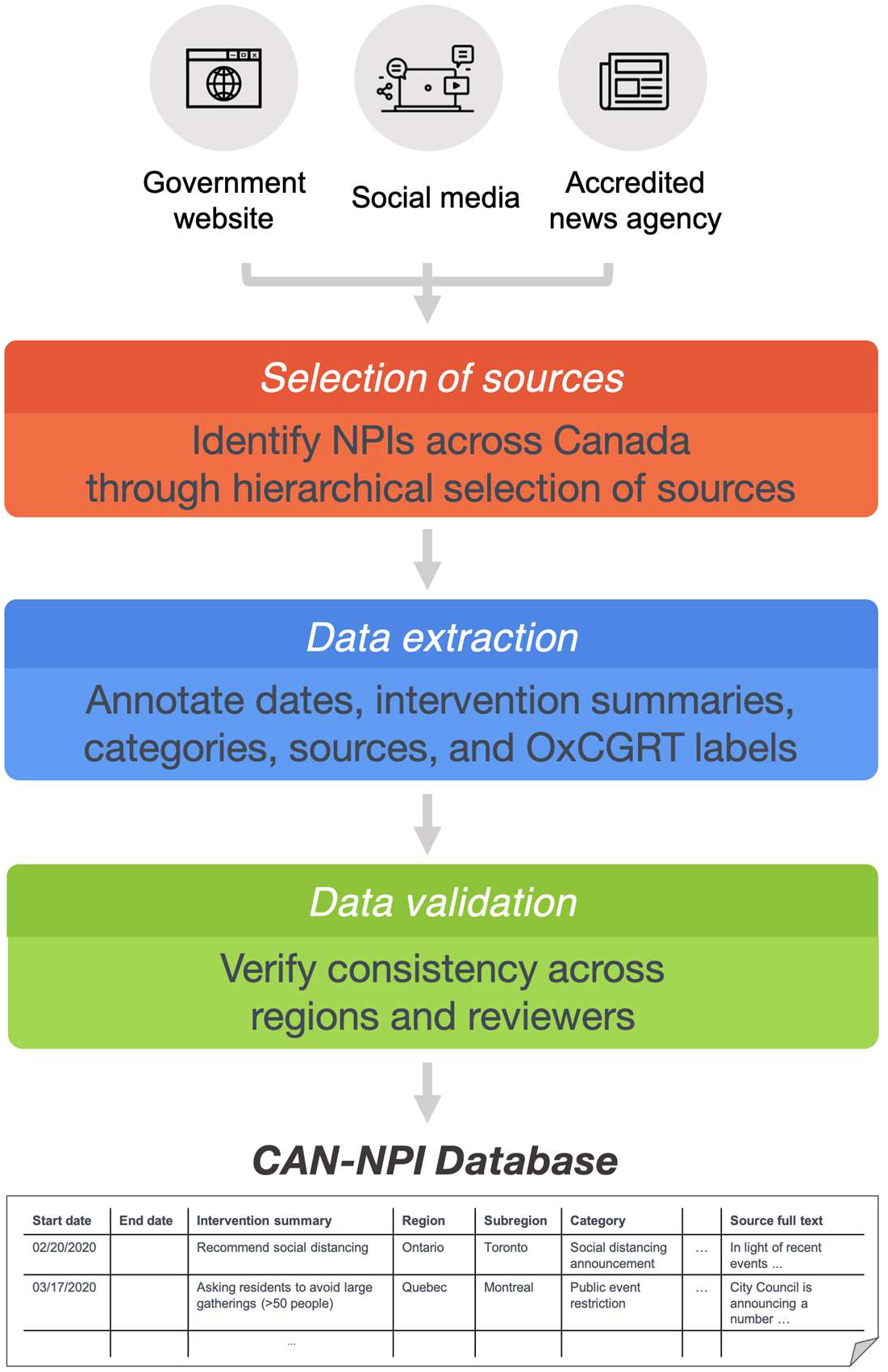
Overview of data collection and validation process for the CAN-NPI dataset.

### Data extraction and NPI classification schema

For each source, after an initial verification check for redundancy with a previous announcement of the same NPI was completed, information was systematically extracted using a consistent format (Supplementary Material Table S2).

Each NPI was labelled with a free-text summary and categorical classifiers describing the nature of the intervention and its target population. Given the shifting nature of the pandemic response, our list of categories was iteratively expanded and adjusted as novel classes of NPIs were identified. If a reviewer was uncertain regarding the appropriate classifier to select, the item was flagged and discussed collectively until consensus was obtained by at least 3 authors.

Our consensus labelling schema contained 63 classes. Additional to these, the interventions could be assigned one of 13 labels aligned with the methodology of the Oxford COVID-19 Government Response Tracker (OxCGRT)^7^. The Oxford group defines 13 interventions in 3 broad categories: decisions related to public gatherings (distancing), financial indicators, and testing and tracing categories. (The OxCGRT group also defines an Oxford Stringency Index using simple scoring of the 7 distancing measures, details of which are provided in Supplementary Material Table S3.)

### Data validation

To ensure data was recorded in a consistent format by each reviewer, we established a streamlined onboarding protocol and a step-wise data-entry process. Standardized data entry forms and skip patterns were also used to minimize data input errors. The data was audited for consistency by evaluating the relative proportions of specific intervention categories between regions. A focused second review of the dataset was also performed by a subset of authors to identify discrepancies and further standardize for consistency and accuracy across reviewers and jurisdictions.

### Dataset Characteristics

We identified information on 1,640 interventions from 899 unique source URLs and 199 unique source organizations across the federal government, all 10 provinces and 3 territories, and the 20 largest CMAs in Canada. This version of the dataset covers interventions starting between January 1st (24 days before the first case was reported in Canada^9^) and March 31st 2020 inclusive, with time being recorded on a daily basis and the median intervention date falling between March 15 and March 24 at a provincial level. The distribution of interventions by daily count over time is shown in **Figure 2A**.

**Figure 2.**
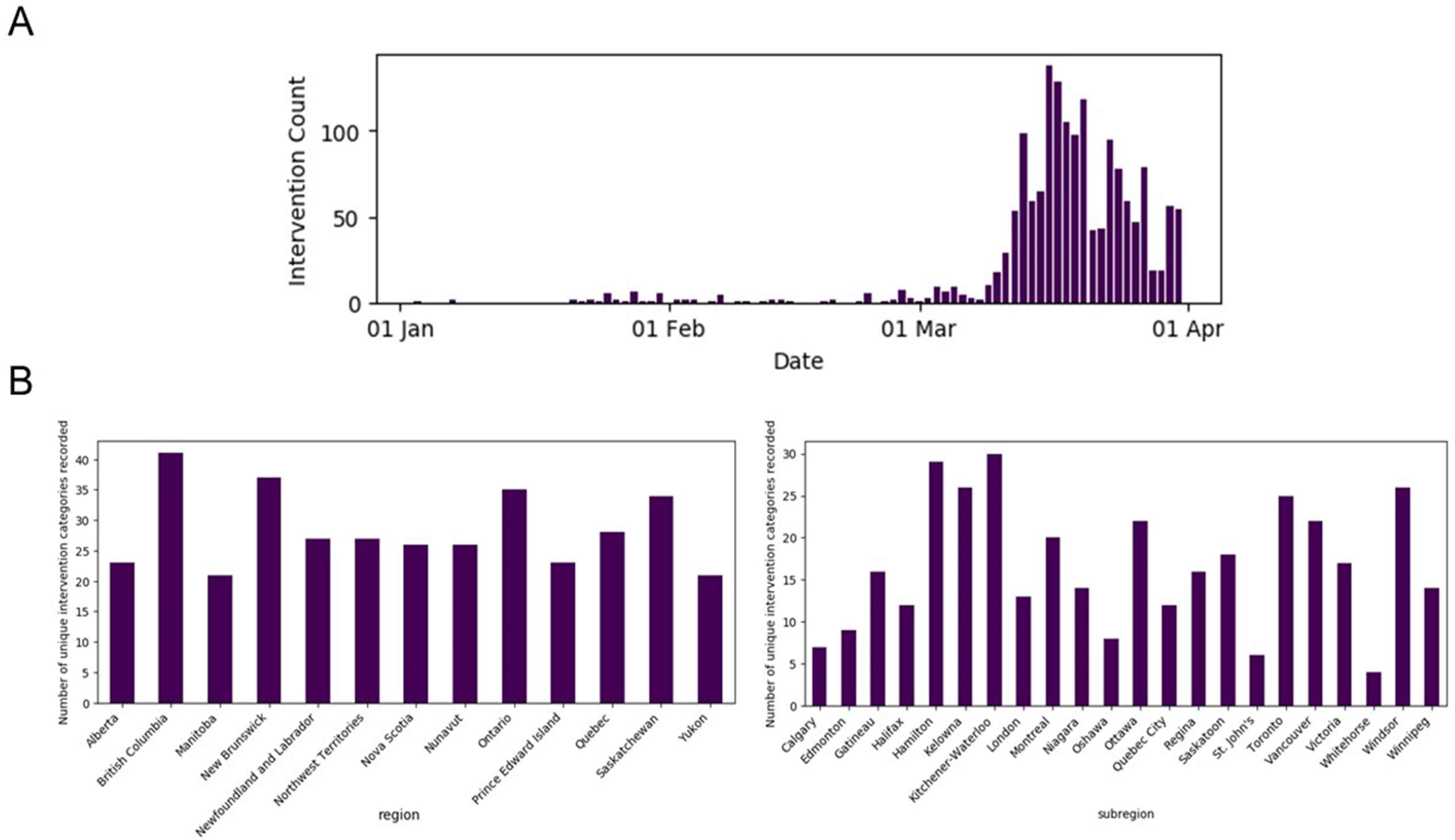
a) Timeline of non-pharmaceutical interventions recorded across Canada in response to the COVID-19 epidemic b) Counts of unique non-pharmaceutical intervention categories recorded for each Canadian region (left) and subregion (right) in response to the COVID-19 epidemic, n=1640

There are 63 categories of intervention in this schema in addition to the 13 OxCGRT interventions. Interventions are also classified based on 13 categories of target populations. The top 15 intervention categories are enumerated in Table 1 with a single example provided for each category. The classes are imbalanced, and most (55%) are classified under the Oxford classification in addition to our categories. The top intervention categories without a corresponding Oxford response category are public announcements, general case announcements, and healthcare facility restrictions. Not all intervention categories are recorded in each jurisdiction. **Figure 2B** shows the variation in the number of unique intervention categories recorded in each region covered by the dataset. Two of the 63 categories were not assigned to an intervention in the first dataset release.

**Table 1:** Top 15 categories of NPIs by count with descriptive examples.

| Intervention Category | Count | Sample Intervention Summary |
| --- | --- | --- |
| Public Announcement | 157 | Recommendation for residents to be vigilant, to refer to Sante Montreal |
| General case announcement | 113 | Announce 9 new cases, total 73 in BC |
| Emergency economic funding | 91 | Government to pay Alberta Energy Regulator industry levy for six months: \$113 million |
| Emergency social services funding | 72 | \$3-million Arts and Culture Resilience Supplement to be administered by the BC Arts Council |
| Non-essential workplace closure | 68 | State of emergency declared: All regulated health services providers will cease operations unless the services to be provided are to address essential health care or an emergency health-care situation. |
| Recreational / Entertainment Facility Closure | 65 | Closure of municipally-run cultural and recreational facilities/programs |
| Social Distancing Announcement | 60 | Government "strongly recommends" companies take travel and distancing measures |
| Healthcare facility restrictions | 57 | Banned visits to hospitals |
| School closure | 55 | Fanshawe College suspends on-site services until further notice |
| Recommended self-isolation | 54 | Recommendation of self-monitoring for returning travellers from conference in Toronto where there was a sick contact |
| Public event/ meeting cancellation or postponement | 53 | All March Break programming, camps and drop-in activities are cancelled |
| Emergency healthcare funding | 41 | Investing \$27.6-Million to support child care for essential workers |
| Government building closure | 40 | Closure of recycling centres until further notice |
| Public park closure | 39 | Oshawa adds off-leash park to list of temporary closures |
| Travel Restriction (Internal) | 37 | Residents asked to use Transit for essential travel only |

### Example Dataset Use

We provide a few examples highlighting the utility of this dataset which uniquely contains longitudinal, geographic, and stringency data on NPIs from across Canada:

#### Provincial and territorial Stringency Index over time

Combining the longitudinal, geographic and strincency features of the dataset, we illustrate the evolution of Oxford Stringency Index for each province over time, which highlights differences in subnational decision-making **(Figure 3)**. For this figure, intervention records that fall into one of the seven categories that qualify for computing the Oxford Stringency Index were used (675 out of 1,640 interventions records). The index was established and tracked daily at the provincial and territorial level. The WHO announcement declaring a global pandemic on March 11th^10^ was selected as a reference date.

**Figure 3:**
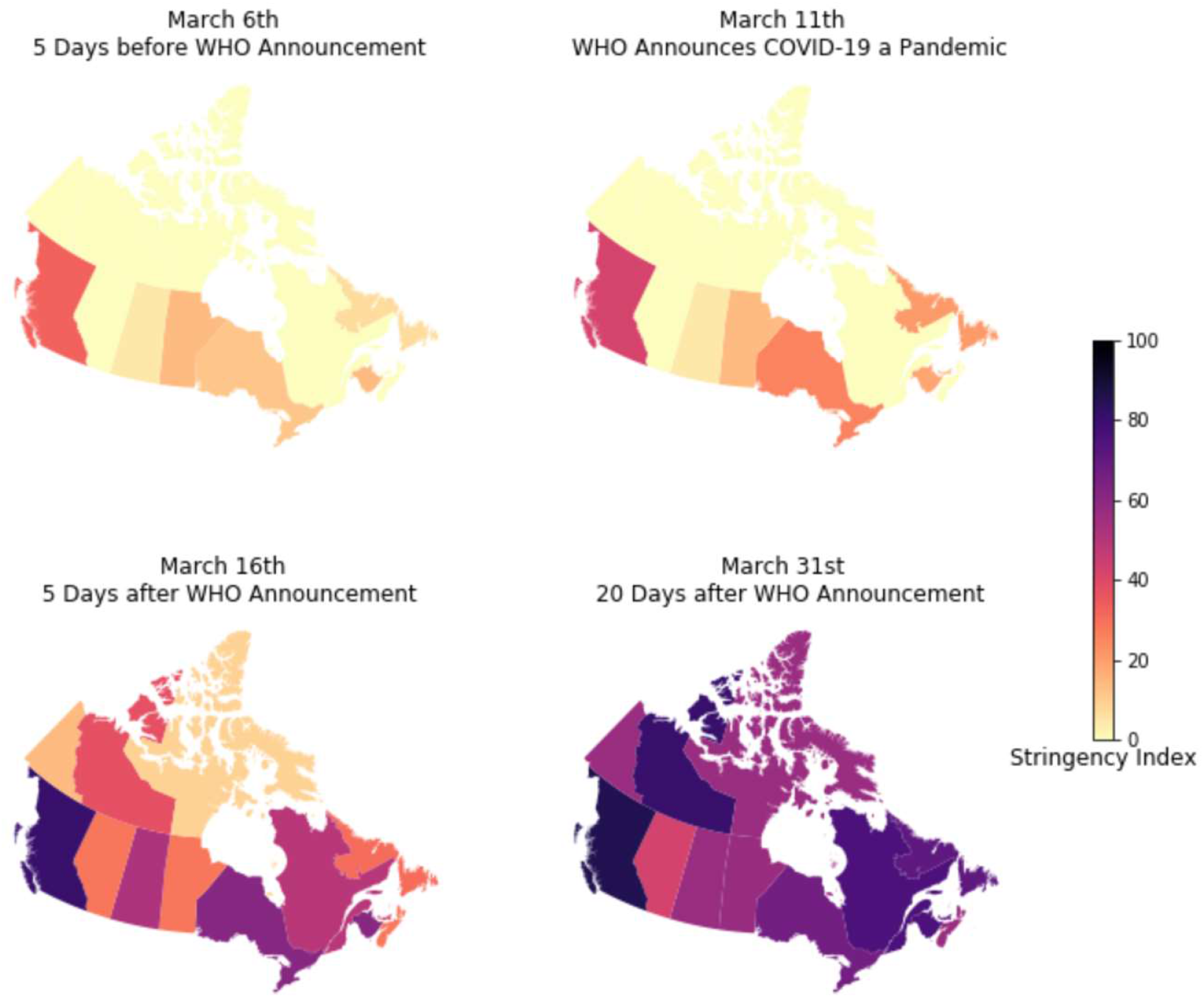
Government Response Stringency Index in Canada by province and territory over time shows temporally increasing stringency with geographic variation (Stringency Index calculated in line with OxCGRT methods (Supplementary Material Table S3)), n=675

#### Time-to-intervention variation across provinces and territories

The jurisdictional and temporal granularity of this dataset allows comparison across provinces and territories of each region’s latency of response to COVID-19. As an example, we show the time-to-intervention for two major NPIs that were instituted in all 13 provinces and territories: declaration of state of emergency (including public health emergency) and school closure. We plot these NPIs on a timeline relative to the first case of and first death due to COVID-19 in each province or territory to visually estimate a time-to-intervention **(Figure 4)**.

**Figure 4:**
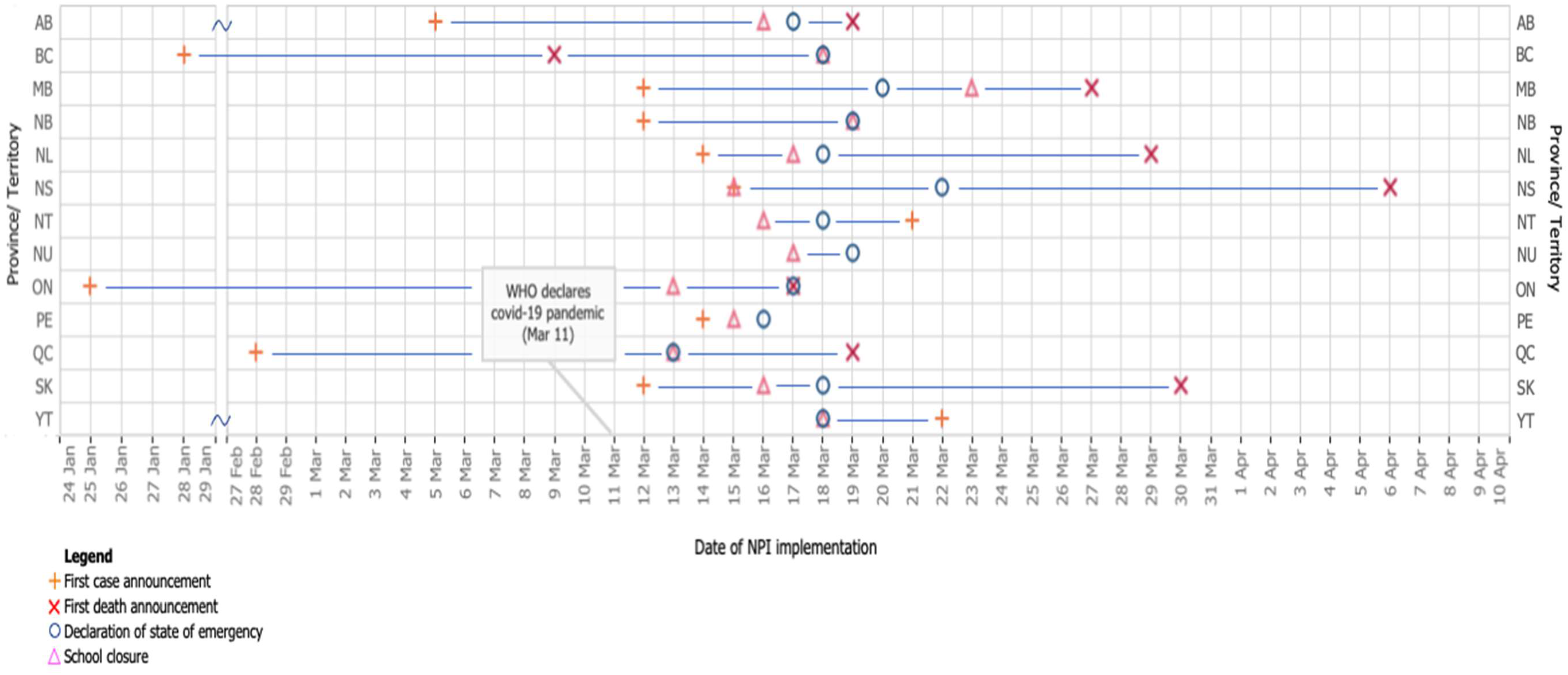
Variation in time-to-intervention by Canadian province / territory for two major NPIs—declaration of state of emergency and school closure— shown relative to two descriptors of the local COVID-19 outbreak, dates of first case and first death in each region.

## Discussion

We have created the first comprehensive dataset of NPIs in response to COVID-19 across Canada that is being made openly available to researchers. Our dataset allows for the comparison of different regions in space and time with regard to their use of NPIs, through both descriptive characterization of the NPIs themselves as well as through time-series based linkage with other data sources (e.g. case and mortality counts, testing, mobility) to enable further analyses of the epidemic’s trajectory in Canada.

The bulk of existing work done on NPIs has been limited to groups simulating the effect of NPIs in mathematical models^2^ or creating region-based collections of NPIs to answer a specific question^5,11,12^. To our knowledge, little work has been done to systematically compile and update NPIs subnationally. While there have been efforts to capture NPIs in the United States at the county level, the data needs to be requested from the user which presents a barrier to access as compared to an open-access download link^13^.

The use of NPIs to combat COVID-19 spread is fundamentally a local issue, in which decision-makers are best guided by data specific to their own locale. While the scope and scale of initiatives such as OxCGRT^7^ or the Assessment Capacities Project (ACAPS) ‘government measures’ dataset^14^ is commendable, these datasets lack uniform granularity in subnational coverage (neither dataset includes subnational entries for Canada at the time of writing). Thus, the emphasis of our work is predominantly its applicability to the Canadian context, while also ensuring compatibility with existing work at the international level.

This dataset has the capacity to enable a wide range of research — both urgently as decision-makers are tasked with understanding and managing Canada’s immediate pandemic response, and in the long-term as retrospective work seeks to understand the nature of this response and how it may be improved for the next pandemic. Given that these public health measures are fundamentally based on the non-occurrence of outcomes (in this case, COVID-19 infections and deaths), such analyses are critical in evaluating the value and appropriateness of various components of the public health response and answering questions about what “should” have been done and when.

Linkages with external sources of high-resolution epidemiologic data^1^, healthcare capacity data, economic impact data, and other subnational NPI datasets (as they become available) will be critical in enabling further research. Our future work involves making these linkages, and providing researchers with streamlined and comprehensive access to high-quality, multi-modal data on the COVID-19 pandemic. We are also seeking to align our data into the global work being done by groups such as OxCRT to allow for robust global analyses. We will continue to update this dataset and others, as well as to offer these analyses, for the duration of the pandemic.

### Limitations

Due to our reliance on public information and releases in creating this dataset, it is possible that interventions that were not publicly announced may have been omitted from the dataset. For example, Nunavut is offering up to $5000 in support for businesses in the territory, but they give no indication of the number of eligible businesses^15^, and thus we could not compute and report a total fiscal value. Similarly, testing policy changes that were not communicated directly to the public through online announcements, such as substantial shifts in practice occurring within public health units and hospitals (e.g. whether or not a testing referral is accepted), may not have been captured in this dataset.

There is inherent variability in how different jurisdictions choose to report and describe their NPIs, as well as the range of information that they choose to include or not include. This may have introduced a degree of variability in our labelling. We have sought to minimize inconsistencies across jurisdictions and reviewers through the aforementioned standardized onboarding process, step-wise data-entry, and a secondary, focused review by a smaller group of reviewers prior to data release. Moreover, as residual variation is likely due to subjective differences rather than error, the open nature of our data allows for end users to suggest future improvements to our dataset or download it to make modifications to suit their specific research needs.

## Conclusion

In just over a month since the declaration of the COVID-19 global pandemic, we have assembled a comprehensive dataset of non-pharmaceutical interventions at the national and subnational level in Canada (which we will continue to update at two-week intervals throughout the pandemic). By capturing data about NPIs at a fine spatio-temporal resolution and presenting it publicly, we have provided researchers, analysts, and the public a means of evaluating policy responses while the course of the pandemic can still be altered. Researchers can now link NPI data to outcome datasets such as case and mortality counts, healthcare resource use, and testing to help determine the most effective and efficient distancing and re-opening strategies. This will be critical if further pandemic waves are to come. Perhaps most importantly, this dataset enables analysis at the provincial, territorial, and municipal levels, where the most impactful decisions will likely be made in Canada. We hope this dataset can act to inform rapid evidence-based policymaking to help flatten the curve, as well as to support retrospective analyses in understanding the spread of COVID-19 in Canada.

## Data Availability

The dataset will be distributed via the website https://howsmyflattening.ca/#/data, as well as on Kaggle (https://www.kaggle.com/howsmyflattening/covid19-challenges) and GitHub (https://github.com/jajsmith/COVID19NonPharmaceuticalInterventions).

## Acknowledgements

*Members of the COVID-19 Canada Open Data Working Group: Non-Pharmaceutical Interventions:* Maya Abdalla, Tasnim Abdalla, Andy Edem Afenu, Kavya Anchuri, Jason Baek, Benjamin Baker, Vishali Balasubramaniam, Isha Berry, Holly Burrows, Yuri Chaban, Joseph Chon, Ryan Daniel, Kristen Dietrich, Vinyas Harish, Sophie Hu, Joseph Jamnik, Mona Khalid, Valerie Kim, Andrew Lam, Haoyue Helena Lan, Liam G. McCoy, Victoria Mintsopoulos, Nykan Mirchi, Ngoc Son Nguyen, Joanna Pineda, Jayoti Rana, Saad Shakeel, Amrit Sampalli, James Saravanamuttu, Jonathan Smith, Jacqui van Warmerdam, Arnold Yeung, Seung Eun Yi, Jennifer Zheng

## Distribution

The dataset will be distributed via the website howsmyflattening.ca, as well as on Kaggle (https://www.kaggle.com/howsmyflattening/covid19-challenges) and GitHub (https://github.com/jajsmith/COVID19NonPharmaceuticalInterventions).

## About Howsmyflattening

Howsmyflattening is a team of physicians, medical students, computational health researchers, designers, engineers and epidemiologists across academia and industry who rapidly gather, integrate, and display data to inform Ontario’s decision-making around the management of COVID-19. This collective will systematically centralize open source data and tackle priority problems while producing data visualizations of challenges related to capacity planning, testing and tracing, regional nuances, and intervention planning. Guided by a team of experts with connections to the COVID-19 decision-making tables, this collaborative will enhance Ontario’s response through cutting-edge data visualizations and triangulation of multiple data sources.

## Legal

This dataset is shared under the Creative Commons CC BY 4.0 License (https://creativecommons.org/licenses/by/4.0/) allowing for sharing and adaptation with credit given. This paper must be cited whenever this dataset is used.

*Icon attributions for Figure 1:*

Website icon by b farias from the Noun Project https://thenounproject.com/search/?q=website&i=1014821 News icon -by Seb Cornelius from the Noun Project (https://thenounproject.com/search/?q=news&i=169028)

Social media icon by Souvik Bhattacharjee from the Noun Project https://thenounproject.com/search/?q=social%20media&i=1631848

## Supplementary Material

**Table S1:** Census Metropolitan Areas Included as Subregions (Population from Statistics Canada, 2016)^15^

| Province | Census Metropolitan Area | Population in 2016 |
| --- | --- | --- |
| Ontario | Toronto | 5,429,524 |
| Quebec | Montreal | 3,519,595 |
| British Columbia | Vancouver | 2,264,823 |
| Alberta | Calgary | 1,237,656 |
| Alberta | Edmonton | 1,062,643 |
| Ontario | Ottawa–Gatineau | 989,657 |
| Manitoba | Winnipeg | 711,925 |
| Quebec | Quebec City | 705,103 |
| Ontario | Hamilton | 693,645 |
| Ontario | Kitchener-Waterloo-Cambridge | 535,154 |
| Ontario | London | 383,437 |
| British Columbia | Victoria | 335,696 |
| Nova Scotia | Halifax | 316,701 |
| Ontario | Oshawa | 308,875 |
| Ontario | Windsor | 287,069 |
| Saskatchewan | Saskatoon | 245,181 |
| Ontario | St. Catharines–Niagara Falls | 229,246 |
| Manitoba | Regina | 214,631 |
| Newfoundland and Labrador | St. John's | 178,427 |
| British Columbia | Kelowna | 151,957 |

**Table S2:** Dataset Schema for NPI Labelling. “Oxford Categories” are adopted from the methodology of the University of Oxford’s Blatnik School of Government Working Paper “Variation in Government Responses to COVID-19”^7^

| Fields | Description | Optional | Schema |
| --- | --- | --- | --- |
| **start_date** | Date of intervention announcement |  | mm/dd/yyyy (Date) |
| **end_date** | Expected end date of intervention | Y | mm/dd/yyyy (Date) |
| **reviewer** | Name of person who added the intervention |  | String |
| **country** | Canada |  | String |
| **region** | Province or All |  | String |
| **subregion** | City, county, provincial health unit, or other | Y | String |
| **intervention_summary** | Manual summary of the intervention |  | String |
| **intervention_category** | Primary category of the intervention |  | *intervention_category* |
| **target_population_category** | Population directly affected |  | *target_population_category* |
| **enforcement_category** | Primary method to enforce intervention compliance | Y | *enforcement_category* |
| **oxford_government_response_category** | Oxford category if applicable | Y | *oxford_government_response_category* |
| **oxford_closure_code** | 0 - No measures, 1 - Recommended measure, 2 - Required measure | Y | *oxford_closure_code* |
| **oxford_public_info_code** | 0 - No campaign, 1 - Campaign | Y | *oxford_public_info_code* |
| **oxford_travel_code** | 0 - No Measures, 1 - Screening, 2 - Quarantine on high-risk regions, 3 - Ban on high-risk regions | Y | *oxford_travel_code* |
| **oxford_geographic_target_code** | 0 - Targeted, 1 - General | Y | *oxford_geographic_target_code* |
| **oxford_fiscal_measure_cad** | New financial measures | Y | Float |
| **oxford_monetary_measure** | New interest rate after change | Y | Float |
| **oxford_testing_code** | 0 - No testing policy<br>1 - Only testing if both a) symptomatic and b) key workers, hospitalized, return from overseas, contact of known case<br>2 - Testing anyone showing COVID-19 symptoms<br>3 - Open public testing for all, including asymptomatic | Y | *oxford_testing_code* |
| **oxford_tracing_code** | 0 - No contact tracing<br>1 - Limiting tracing; not for all cases<br>2 - Comprehensive tracing; done for all cases | Y | *oxford_tracing_code* |
| **source_url** | Website URL of source containing intervention announcement |  | URL |
| **source_organization** | Primary announcing or responsible organization | Y | String |
| **source_organization_2** | Secondary or joint announcing or responsible organization | Y | String |
| **source_category** | Government Website, Accredited News Agency, or Social |  | *source_category* |
|  | Media |  |  |
| **source_title** | Title of source containing intervention announcement | Y | String |
| **source_full_text** | Full text of source containing intervention announcement | Y | String |

The Oxford Stringency Index (OSI) is a simple score calculated using a sum of the first 7 of 13 classes (S1 to S7).^5^ For S5, a score of 0 or 1 is given, and for the other six types, a score between 0 and 3 is given depending on the strictness of the government measure. Each score is then scaled to take a value between 0 and 100 and the seven scores are averaged to get the composite stringency index. For instance, a generalized, recommended school closing announcement would correspond to a score of 2 out of 3, resulting in the final score of 67. This index is indicative of the number and the strictness of different government interventions, but should not be interpreted as effectiveness. Our dataset allows us to compute this index for each province and territory to better understand the evolution of government response across Canada over time.

**Table S3:** (a) Oxford intervention categories for calculation of the Oxford Stringency Index adopted from “Variation in Government Responses to COVID-19”^5^

| Oxford Category ID | Oxford Category Name | Coding Scheme |
| --- | --- | --- |
| S1 | School Closing | Ordinal (0-2) + Binary for geographic scope |
| S2 | Workplace Closing | Ordinal (0-2) + Binary for geographic scope |
| S3 | Cancel public events | Ordinal (0-2) + Binary for geographic scope |
| S4 | Close public transport | Ordinal (0-2) + Binary for geographic scope |
| S5 | Public info campaigns | Binary + Binary for geographic scope |
| S6 | Restrictions on internal movement | Ordinal (0-2) + Binary for geographic scope |
| S7 | International travel controls | Ordinal (0-2) + Binary for geographic scope |
| S8 | Fiscal measures | Numerical (CAD) |
| S9 | Monetary measures | Numerical for interest rate (%) |
| S10 | Emergency investment in healthcare | Numerical (CAD) |
| S11 | Investment in vaccines | Numerical (CAD) |
| S12 | Testing policy | Ordinal (0-2) |
| S13 | Contact tracing | Ordinal (0-2) |

**Table S4:**
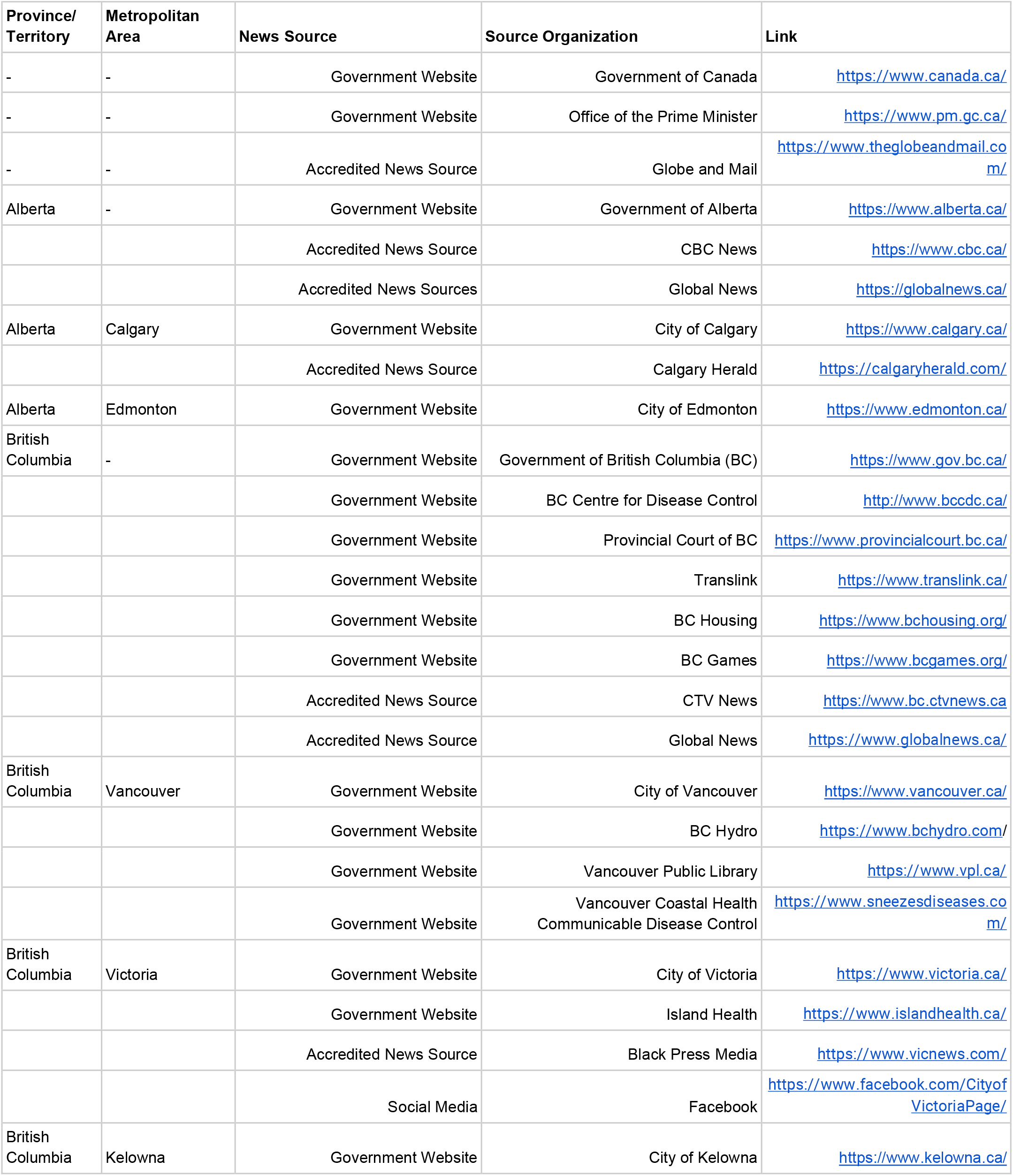
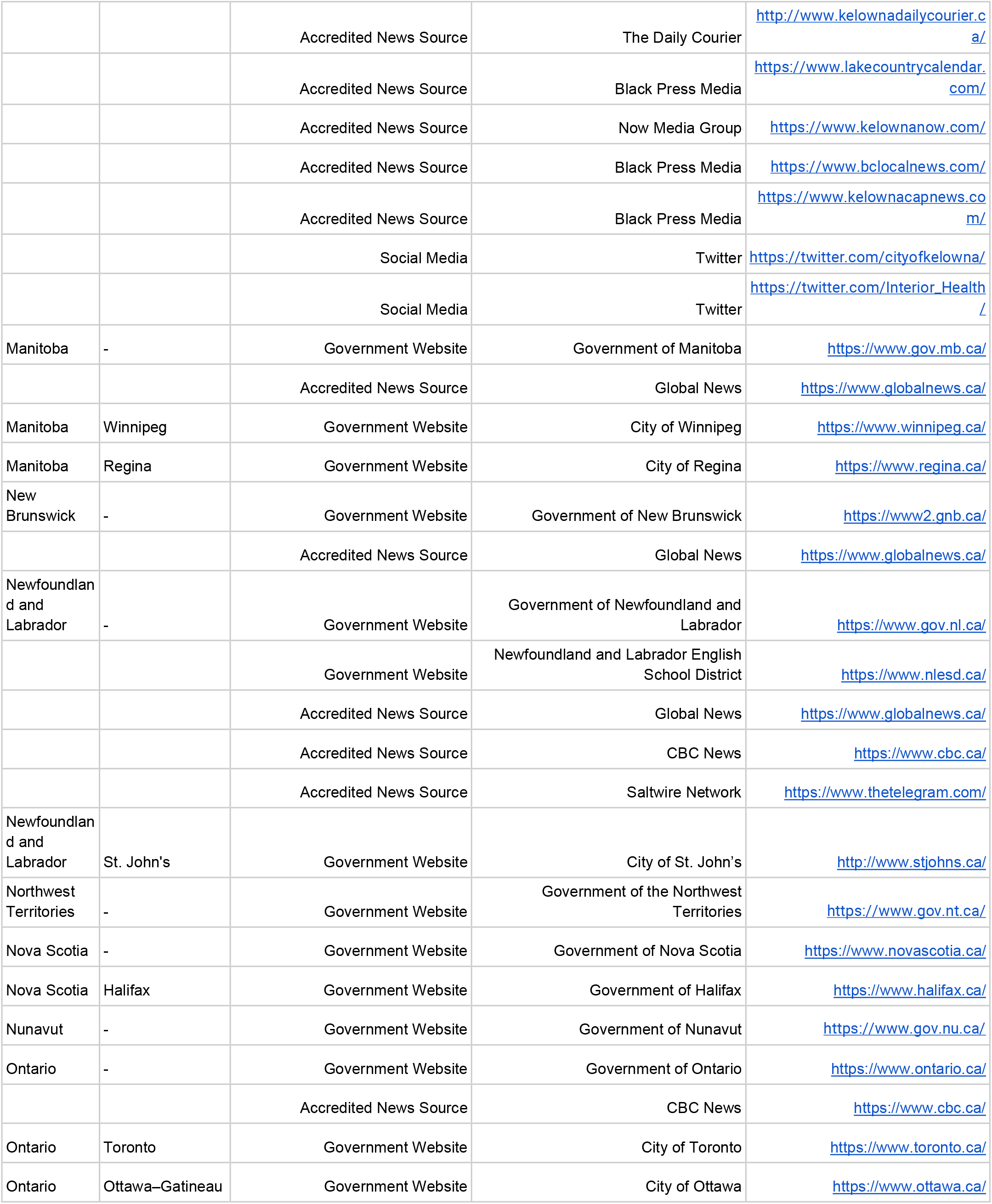
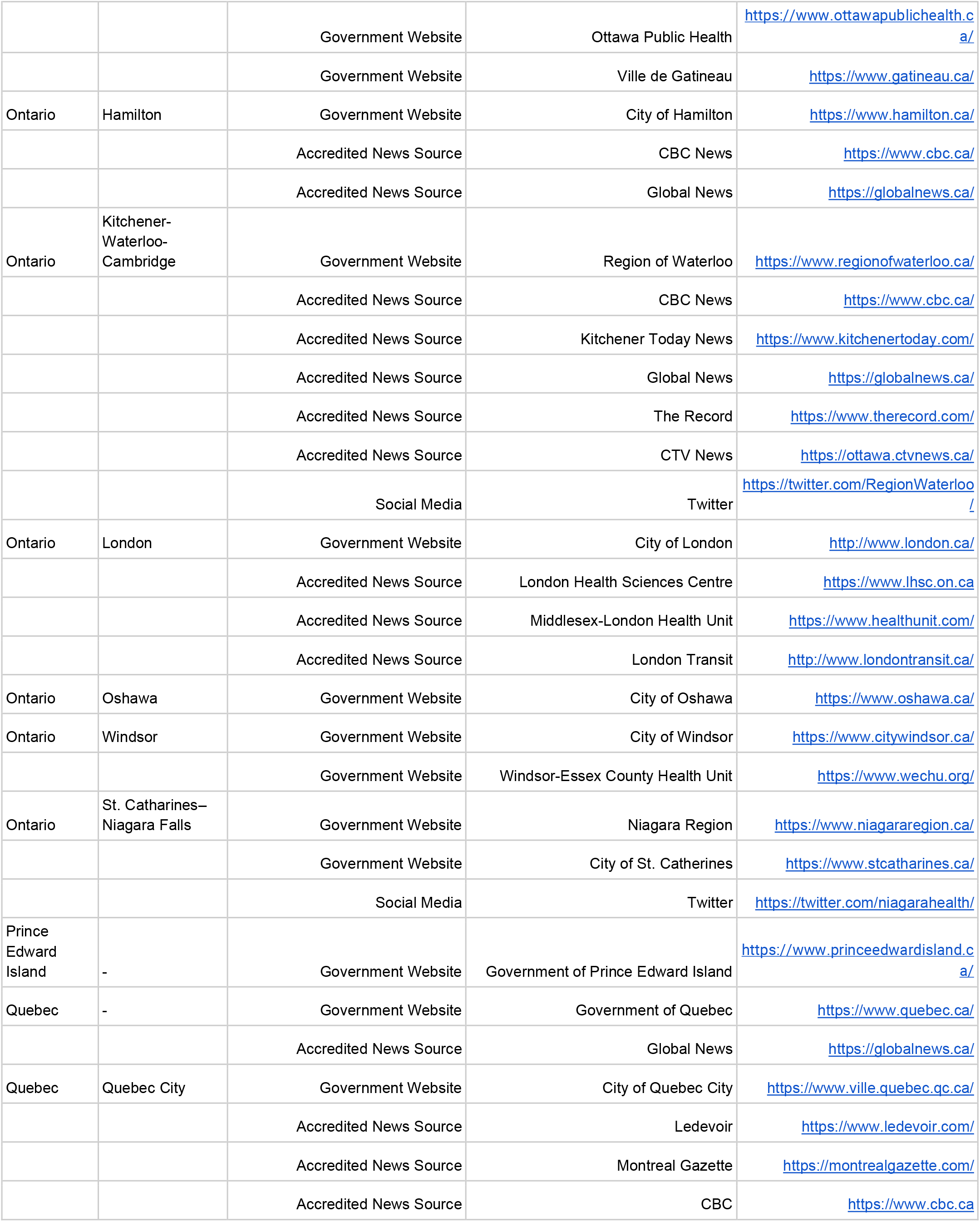
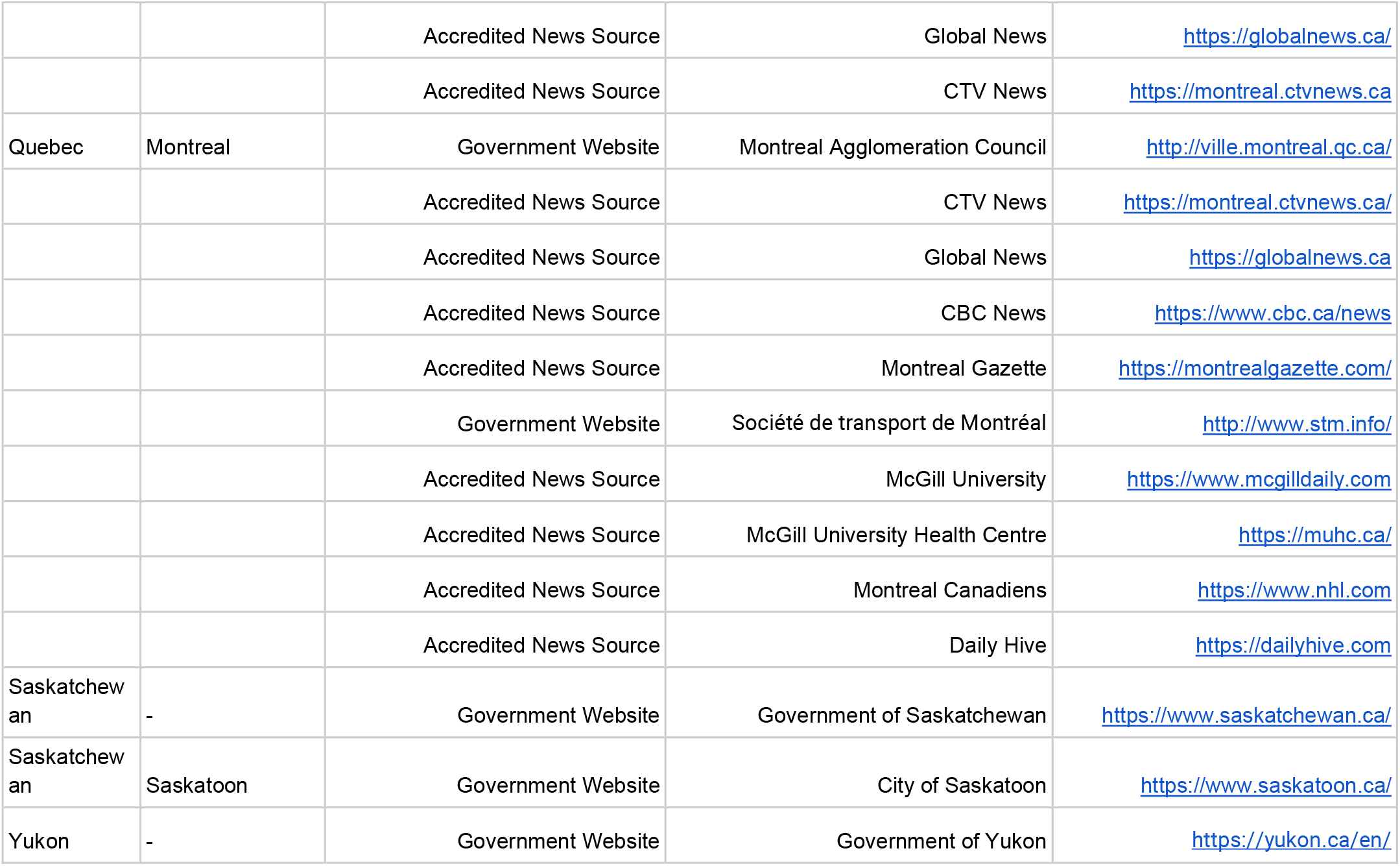
Federal, provincial, territorial and regional sources by source type, organization and link

